# Association of socioeconomic status with arterial stiffness in older African American and White Adults: The ARIC Study Cohort

**DOI:** 10.1101/2022.05.05.22274728

**Authors:** T.A. Spikes, A.B. Alam, T.T. Lewis, B.G. Windham, A. Kucharska-Newton, A. Alonso

**Author notes:** Corresponding Author: Telisa Spikes, Nell Hodgson Woodruff School of Nursing, 1520 Clifton Rd. Office# 428 Atlanta, GA 30322.

## Abstract

**Background:** The contributions of traditional CVD risk factors do not fully explain the racial and socioeconomic disparities in arterial stiffness. The extent to which the association of socioeconomic status (SES) with subclinical CVD is different for older African American and White adults is unclear.

**Methods and Results:** Cross-section associations of individual SES [education and income] with carotid femoral pulse wave velocity (cfPWV), a subclinical marker of arterial stiffness, and modification of this relationship by race were examined among 3,342 participants of the Atherosclerosis Risk in Communities (ARIC) Study (age=75±4.9 years, 64% female, 23% African American, free of CVD in 2011-2013) using multivariable linear regression. Post-graduate education compared to less than high school, was associated with lower cfPWV (less stiffness) in African Americans (*β* = −1.28 m/s; 95% CI, −1.97, −0.59), but not in Whites (*β =* −0.69 m/s; 95% CI, −1.39, 0.01). Income ≥$50K as compared to <$25K, was associated with lower cfPWV both in African Americans (*β* = −0.82 m/s; 95% CI, −1.42, −0.22) and Whites (*β* = −0.76 m/s; 95% CI, −1.19, −0.32). However, interaction of race with individual measures of SES on cfPWV in African American and White adults were not statistically significant (p-value for interaction >0.05).

**Conclusions:** Higher SES was cross-sectionally associated with lower arterial stiffness in this cohort; the data did not support differences by race. Prospective studies of SES and cfPWV among a racially socioeconomically diverse cohort merit further study.

## Introduction

Despite overall decreases in mortality from cardiovascular disease (CVD) observed in recent decades,^1,2^ important disparities exist that are patterned by race and socioeconomic status (SES).^3^ Compared to White populations and those of high SES, Black populations and persons of low SES experience a greater burden of traditional CVD risk factors and higher blood pressure levels that may also contribute to greater arterial stiffness, a subclinical marker of CVD.^4,5^ However, the contribution of traditional CVD risk factors does not fully explain the racial and socioeconomic differences in arterial stiffness.^6^ A growing body of evidence suggests that racial minority groups experience fewer health gains from attaining a higher socioeconomic position compared to White individuals, which may partially explain the enduring racial and ethnic disparities underlying CVD.^7–10^ Identifying racial and socioeconomic disparities at the subclinical level has important implications for the prevention of overt CVD and target organ damage.^11^

Arterial stiffness, estimated most often from measures of aortic pulse wave velocity (PWV),^12^ is acknowledged as a predictor of future CVD and all-cause mortality independently of other cardiovascular risk factors.^4,13^ Despite the well-established SES-health gradient,^14^ inconsistent SES patterns as well as differential vulnerabilities to arterial stiffness have been observed among higher versus lower SES Black adults whereas these patterns were undetected in White populations.^15,16,^ These findings may be the result of the Marginalization-related Diminished Returns (MDRs) theory, which posits that the effects of SES indicators tend to be weaker on economic, behavioral, and health outcomes for the members of socially marginalized versus members of the socially priviledged groups.^9,17^ Furthermore, prior studies have found a flatter SES-health association for Black versus White populations that are attributed to structural factors (i.e. poverty, historical and contemporary forms of racism and discrimination)^18,19^ resulting in racial health disparities that are most pronounced at the highest levels of SES.^17,20,21^ Due to these issues, researchers have argued that it is necessary to not only examine the main effects by which SES operates, but also test for SES-race interactions as they reflect the intersection of different systems of constraint and opportunity with respect to health.^19,21,22^

The extent to whether associations of education and income with arterial stifness are weaker or differential in older African American and White adults remains unclear. In the present study, we examined the cross-sectional associations of individual measures of SES—education and income—with PWV, and race as an effect modifier between SES and PWV, in a cohort of older African American and White adults. We hypothesized that the associations between individual measures of SES and arterial stiffness as well as the effect of higher education and income would be weaker for African American versus White adults.

## Methods

### Study Population

The Atherosclerosis Risk In Communities (ARIC) Study is an ongoing community-based, longitudinal prospective cohort of 15,792 men and women, age 45-64 years at baseline, recruited from four US communities: Forsyth County, NC (African Americans and Whites); Jackson, MS (all African Americans); selected suburbs of Minneapolis, MN; and Washington County, MD. Recruitment began in 1987 with baseline screening occurring through 1989. Subsequent follow-up visits were conducted in 1990-1992 (visit 2), 1993-1995 (visit 3), 1996-1998 (visit 4), 2011-2013 (visit 5), 2016-2017 (visit 6), and 2018-2019 (visit 7). Additional information about the initial study design and objectives can be found elsewhere.^23,24^

Pulse wave velocity (PWV) was measured at visit 5 (2011-2013) among 6538 participants, with 6496 participants with PWV measurements. Participants were excluded if they had missing information on SES variables [educational attainment (*n*=11), median household income (*n*=664)]; a history of prevalent coronary heart disease (*n*=645), heart failure (*n*=210), or stroke (*n*=153); if they had any of the following: body mass index (BMI) ≥ 40kg/m^2^ (*n*=143); major arrhythmias (Minnesota code 8-1-3, 8-3-1, and 8-3-2) (*n*=146), Minnesota code 8-1-2 with evidence of low quality PWV waveforms (*n*=153), aortic stenosis (*n*=28), aortic regurgitation (*n*=24), aortic aneurysm (*n*=82), self-reported aortic revascularization surgery (*n*=54), and peripheral revascularization (*n*=28) due to interference with PWV measurements;^25^ or missing covariates of interest: BMI (*n*=267), systolic (*n*=35) and diastolic (*n*=35) blood pressure, and current smoking status (*n*=423). Further, participants who self-identified as non-African American or non-White (*n*=18) were excluded from these analyses. Participants who self-identified as African American from the Maryland (*n*=15) and Minnesota (*n*=9) sites were also excluded due to the small sample size, leaving a final analytic sample of 3,362 participants (Figure 1. Flow diagram). Institutional Review Boards at each of the four sites approved the study, and written informed consent was obtained from all participants at each study visit.

**Figure 1.**
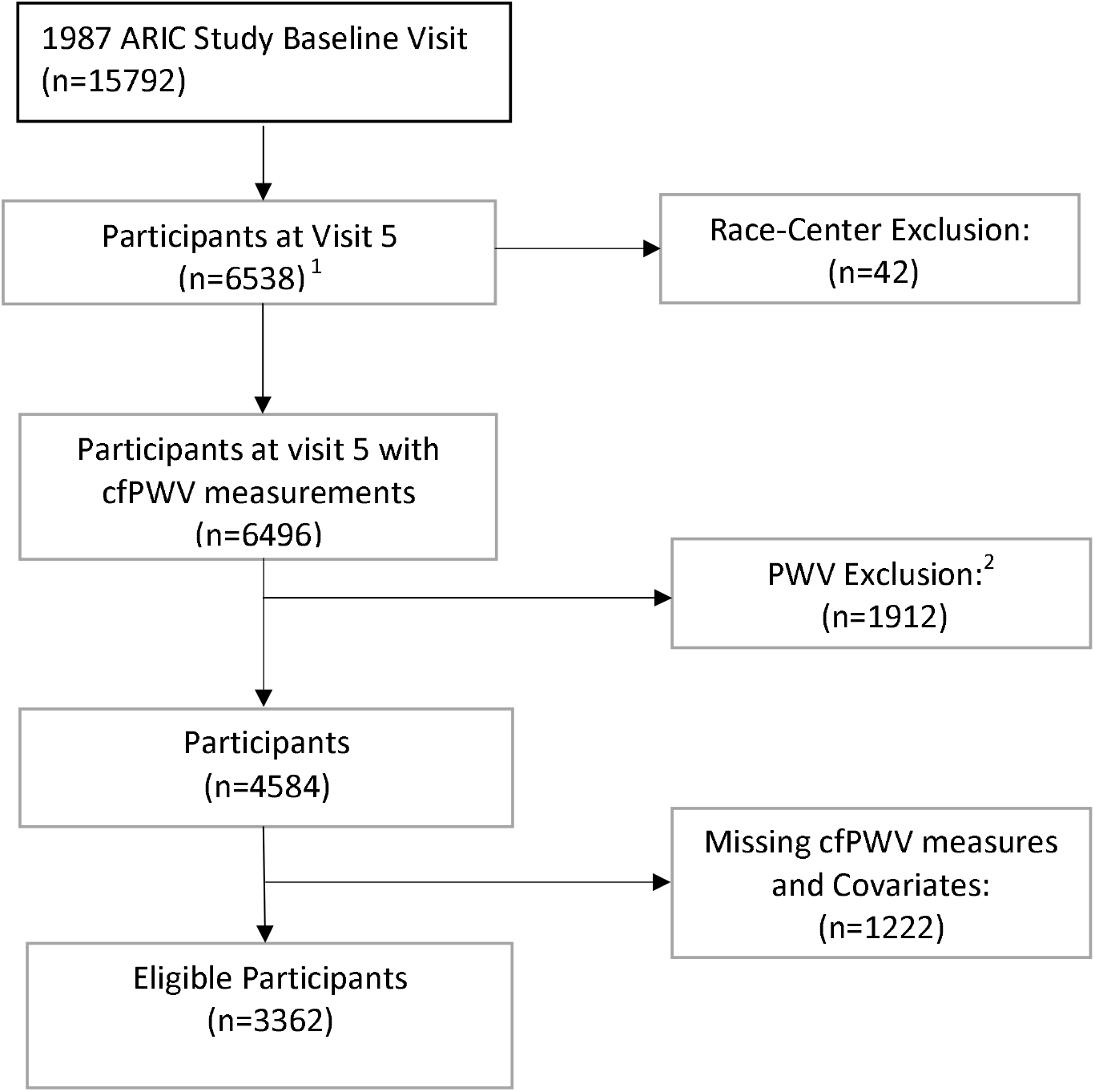
Process of Selection of Study Population Abbreviations: ARIC, Atherosclerosis Risk in Communities Study. ^1^Participants in primary analysis; ^2^Predefined factors that might affect the validity of PWV measurements.

### Measures

#### Socioeconomic Status

Two individual socioeconomic status indicators were included in this study: educational attainment and total combined family income. Educational attainment was characterized as highest level of education reported at visit 1 (1987-1989) and categorized into 4 categories: Less than High School (HS) (*n*=389), HS (HS/GED, *n*=1123), Some College and above (Vocational school and college, n=1330) and Post-Graduate (Graduate school, *n*=520). Total combined family income was self-reported at visit 5 (2011-2013). Total combined family income, self-reported at visit 5, was categorized as: <$25,000 (*n*=865); $25,000-<$50,000 (*n*=1106); and $50,000 and above (*n*=1391).

#### Outcome

Arterial stiffness was estimated from measurements of carotid femoral PWV (cfPWV) performed during visit 5 (2011-2013) by trained technicians using an automated waveform analyzer VP-1000 Plus (Omron, Kyoto, Japan) following a standardized protocol, with faster cfPWV indicating higher arterial stiffness.^26^ Participants were supine for 5-10 minutes prior to the measurement. Carotid and femoral arterial pressure waveforms were acquired by applanation tonometry sensors on the left common carotid artery and left common femoral artery. Pulse wave velocity was calculated as the distance between the suprasternal notch to carotid minus the carotid to femoral distance divided by transit time.^25^ Distance for cfPWV was measured with a segmometer (Rosscraft, Surrey, Canada). For the purposes of these analyses, cfPWV was modeled as a continuous variable in meters/second.

Quality control procedures regarding PWV assessment included central training and recertification, quarterly equipment calibration, and ongoing quality control reviews on a random sample of 40 records per month stratified by center with feedback provided to technicians.^27^ Approximately 78% of records were considered optimal quality, 17% were good quality, 3% were acceptable, and none were poor or unacceptable.^27^

#### Covariates

Other than sex, race, and educational attainment, which were assessed at ARIC ^24^study baseline, all covaraites considered in this analysis were ascertained at visit 5 (2011-2013). Age, race, sex, current alcohol consumption, and smoking status were self-reported. Body mass index (BMI) was calculated as measured weight in kilograms divided by height in meters squared. Systolic blood pressure (SBP) and diastolic blood pressures (DBP) were obtained by taking the averages of the second and third of three blood pressure measurements. HTN was defined as SBP ≥ 140mm Hg and DBP ≥ 90mm Hg or antihypertensive medication use. Diabetes mellitus was defined as: a fasting blood glucose level ≥126 mg/dL, non-fasting blood glucose ≥200 mg/dL, use of hypoglycemic medication(s), or a self-reported physician’s diagnosis. Circulating total cholesterol and high-density lipoprotein (HDL-C) were measured in collected plasma samples using standard procedures.^28^

### Statistical Analysis

Characteristics of the study population are presented as means for continuous variables and proportions for categorical variables overall and by educational attainment, income and race. Associations of individual SES measures with cfPWV were examined separately in minimally and fully adjusted models, both in the overall cohort and separately by race using multivariable linear regression analyses. For the overall analyses, *model 1* adjusted for age, sex (male or female), race/ethnicity (African American or White), and ARIC race-center variable; *model 2* added HDL-C, total cholesterol, BMI, smoking (yes or no), current alcohol use (yes or no), lipid lowering medication use (yes or no), and diabetes (yes or no); and *model 3* added SBP, DBP and BP medication use (yes or no). To test whether the association between SES indicators and arterial stiffness differed by race, we included interaction terms between educational attainment and race and income and race. Interaction effects of race and individual SES measures on cfPWV were tested for significance based on the Wald Chi-square statistic. Statistical analyses were performed using SAS 9.4 (Cary, NC) and STATA, version 16 (StataCorp LP, College Station, Texas).

## Results

### Participant Characteristics

Participant characteristics, by educational attainment are shown in Table 1. Participants with lower education were slightly older, were less likely to be alcohol drinkers, and had a worse overall cardiovascular risk profile in both Whites and African American adults. Characteristics of the cohort by income and race are shown in Table 2. Participants with lower income were more likely to be older and female, less likely to drink alcohol, likely to be current smokers, and had a higher prevalence of cardiovascular risk factors.

### Associations of educational attainment and median household income with cfPWV

The association between education, income, and cfPWV for the overall sample is shown in Table 3. Compared to less than a HS education, a post-graduate education was associated with lower cfPWV in the minimally adjusted model 1, (β=−0.94 m/s, *95*% CI=−1.44, −0.44). Subsequent adjustment for CV risk factors in model 2 attenuated this association yet remained significant (β=−0.63 m/s, *95*% CI=−1.14, −0.12). Following adjustment for BP and BP medications in model 3 resulted in attenuation of this association becoming no longer statistically significant. High income was inversely associated with cfPWV. Compared to participants with income <$25,000, those with income of $25,000-<$50,000 and ≥$50,000 had lower cfPWV [-0.56 m/s (95% CI=−0.91, −0.22) and −0.82 m/s (*95*% CI= −1.16, −0.47), respectively]. Following adjustments for CV risk factors in model 2 and additionally for BP and BP medications in the fully adjusted model 3, these associations remained unchanged among those earning $25,000-<$50,000 and slightly attenuated but significant for those earning ≥$50,000. Race-stratified models for African American and White adults are shown in Supplementary Tables 1 and 2. Post-graduate vs. less than high school education was associated with a −1.29 m/s (*95*% CI= −1.98, −0.60) lower cfPWV in the minimally adjusted model among African American adults. These associations were reduced but remained significant after adjusting for CV risk factors (model 2) and BP and BP medications (model 3). In contrast, post-graduate versus less than high school education had a non-significant and weaker influence on cfPWV in White adults (Table 5; −0.68 m/s, *95*% CI= −1.38, 0.01). Income of $25,000-<$50,000 and ≥$50,000 was associated with a lower cfPWV in the minimally adjusted models for African American participants [(−0.90 m/s (*95*% CI= −1.49, −0.31) and (−0.86 m/s (*95*% CI= −1.46, −0.27)] m/s, respectively (Supplemenatry Table 1). In the fully adjusted model, associations for both income categories were attenuated and no longer significant for those earning ≥$50,000, yet remained significant among those earning $25,000-<$50,000 (−0.69 m/s (95% CI= −1.26, −0.13). The analyses between income and cfPWV for White participants earning ≥$50,000 versus those earning <$25,000 had −0.74 m/s (*95*% CI= −1.17, −0.31) lower cfPWV (Supplemenatry Table 2; model 1). After adjustment for cardiovascular risk factors, these associations were attenuated but remained significant. (Supplementary Table 2; Models 2 and 3). Although stratified results suggested associations might differ by race, formal tests of interaction between race and SES measures of educational attainment and income for the association with cfPWV were not statistically significant (p>.05) (Supplementary Table 3).

## Discussion

In this cross-sectional cohort study of 3,362 older African American and White adults free of CVD, we found inverse associations of income and educational attainment with arterial stiffness as measured by cfPWV. Counter to our hypothesis, the association between higher education and lower cfPWV was stronger in African American adults versus White adults; however, tests of interactions between education and race as well as income and race on arterial stiffness were not statistically significant, failing to provide evidence for effect modification by race.

Findings from the current study offer some key contributions to an existing body of literature that seeks to better understand the relationship between measures of SES and disparate CVD related health outcomes in older African American and White adults. First, our results are contrary to findings from an earlier analysis of the ARIC cohort using beta stiffness index derived from brachial blood pressure and echo-tracked systolic and diastolic carotid arterial diameters, which found that African American persons with a HS or above education compared to those with less than a HS education had greater carotid stiffness.^16^ A recent analysis examining the distribution of arterial stiffness by race and SES also found that higher (versus lower) SES African American adults had greater arterial stiffness which was not observed in White men or women.^15^ Our findings were in the expected direction of the well-established SES-health gradient for both educational attainment and income.^14,29^ In fact, the effect of higher education relative to income was stronger among African American adults than among White adults, thus contradicting the MDRs hypothesis that the health gains would be smaller from similar increases in SES resources for African American relative to White adults.^7,17^ Similar to a prior analysis using the Health and Retirement Study data examining the MDRs of education against self-rated health, BMI, and depressive symptoms, no differences were found for the protective effects of higher education against self-rated health and depressive symptoms for Black or White adults.^30^ One potential explanation for our findings is that the MDRs hypothesis has a larger impact at the higher end of the SES distribution. Recent findings from Spikes et. al in a cohort of African American women from a wide range of SES backgrounds only observed differences in daytime SBP among women making less than $35,000 compared to women earning more than $35,000, with very similar BPs between women earning, $35,000-$49,999, $50,000-$74,999, and more than $75,000.^31^ The limited number of African American participants with a high income and/or a post-graduate education in the ARIC population could limit our ability to observe this pattern. Additionally, our findings may be evidence of resource substitution which posits that individulas who are marginalized within society benefit more from each year of education compared to socially advantaged groups due to limited access to other resources (i.e. power, authority, earnings) and are more dependent upon resources that are available to them.^32^ Further studies are needed to efficiently compare larger racially and regionally diverse populations with a wider range of socioeconomic profiles to characterize the effects of SES resources between African American and White adults.^33^

Similar to a prior analysis of the ARIC cohort,^16,34^ we also did not observe a statistically significant association between education and arterial stiffness for White participants. Among the education levels, a post-graduate education was inversely associated with arterial stiffness, albeit the association was very small. Similar to education, we also observed the expected SES-health association between higher income and lower arterial stiffness. One potential factor that may underlie and explain the influence of income having a stronger effect on arterial stiffness relative to education includes the ability for White populations to leverage upon and gain access to other resources such as higher wage paying jobs, which may be more impactful and protective against poor health.^32,34^ Additionally, potential mediators of education quality and differential pay based on neighborhood/region that affect income and health, could be influential and may mitigate effects of lower education on arterial stiffness in White populations, however further study is warranted to examine these relationships.^34,35^ Future studies including social contextual measures are needed to prospectively evaluate and test these associations in more detail.

Our study has several strengths. This study was performed using data from a large and well-characterized community-based cohort of older African American and White adults free of CVD that represent different geographic regions of the United States. We took advantage of the availability of cfPWV data, which is recognized as the gold standard measure of arterial stiffness, a robust and independent predictor of CVD.^36, 37^ A significant limitation is that our study had a relatively small number of African American participants, almost all of whom were from one field center, and a limited number of White participants in the lowest and African American participants in the highest income categories, impacting our ability to conduct meaningful subgroup analyses or fully disentangle race versus regional influences. Additionally, we were unable to disentagle the effects of race and education specifically for the southern geographic region due to schools being segregated, thus making comparisons ineffectual. Second, our study was cross-sectional, therefore, temporal ordering could not be established. SES is a dynamic and complex measure that extends beyond traditional measures of education and income;^38,39^ however other measures of individual SES such as wealth are not available in this cohort. Lastly, these findings may not be generalizable to other ethnic minority groups or younger adults.

## Conclusions

In conclusion, higher income was associated with lower arterial stiffness in the overall cohort and within race strata. Additionally, higher education among African American adults was associated with lower arterial stiffness. There was no evidence of diminished returns in African American participants. Further detailed examinations into social contextual and cultural contributions of subclinical CVD merits additional study in larger populations representing regional as well as racial and ethnic diversity.

## Data Availability

All data produced in the present work are contained in the manuscript

## Acknowledgments

The authors thank the staff and participants of the ARIC study for their important contributions.

