## Supplementary material for "Association of socioeconomic status with arterial stiffness in older African American and White Adults: The ARIC Study Cohort": Tables 1, 2, 3

**Table 1. Characteristics of study participants by Education and Race, ARIC Study, 2011-2013**

|  | **African American (N = 776)** | | | | | | | | |  | | **White (N = 2573)** | | | | | | |
| --- | --- | --- | --- | --- | --- | --- | --- | --- | --- | --- | --- | --- | --- | --- | --- | --- | --- | --- |
| M(SD) or N[%] | <HS  191 [24] | HS  173 [22] | Some college or college  239 [30] | | | | | | Post-graduate  186 [24] |  | | <HS  198 [8] | HS  950 [37] | | Some college or college  1091 [42] | | Post-  graduate  334 [13] | |
| Age, yrs | 75 (5) | 74 (5) | | 73 (4.3) | | | 74 (4.7) | | | |  | 78 (5.5) | 76 (5.1) | | 76 (5.3) | | | 75 (5.1) |
| **Sex** |  |  | | | | | |  | | |  |  |  | |  | |  | |
| Females | 136 [71] | 129 [75] | 152 [64] | | | | | 135 [73] | | |  | 132 [67] | 658 [69] | | 664 [61] | | | 130 [39] |
| **Site** |  |  | | | | | |  | | |  |  | |  | | | |  |
| Forsyth Co., NC | 6 [3] | 9 [5] | 24 [10] | | | | | 6 [3] | | |  | 24 [12] | 217 [23] | | 284 [26] | | | 104 [31] |
| Jackson, MS | 185 [97] | 164 [95] | 215 [90] | | | | | 180 [97] | | |  | - | - | | - | | | - |
| Minneapolis suburbs, MN | - | - | - | | | | | - | | |  | 25 [13] | 318 [33] | | 560 [51] | | | 149 [45] |
| Washington Co., MD | - | - | - | | | | | - | | |  | 149 [75] | 415 [44] | | 247 [23] | | | 81 [24] |
| **Income** |  |  | | |  | | | | | |  |  | |  | | |  | |
| <$25K | 156 [82] | 117 [68] | 104 [44] | | | | | | 19 [10] | |  | 105 [53] | 220 [23] | | 134 [12] | | | 10 [3] |
| $25K-<$50K | 23 [12] | 44 [25] | 70 [29] | | | | | | 59 [32] | |  | 65 [33] | 417 [44] | | 368 [34] | | | 60 [18] |
| ≥$50K | 12 [6] | 12 [7] | 65 [27] | | | | | | 108 [58] | |  | 28 [14] | 313 [33] | | 589 [54] | | | 264 [79] |
| **Lifestyle** |  |  |  | | |  | | | | |  |  |  | |  | | |  |
| Alcohol drinker | 27 [14] | 25 [15] | 57 [24] | | | | | | 68 [37] | |  | 58 [29] | 491 [52] | | 767 [70] | | | 255 [76] |
| Current smoker | 19 [10] | 11 [6] | 14 [6] | | | | | | 4 [2] | |  | 12 [6] | 57 [6] | | 67 [6] | | | 13 [4] |
| **CV risk factors** |  |  |  | | | | | |  | |  |  |  | |  | | |  |
| Diabetes | 86 [45] | 79 [46] | 93 [40] | | | | | | 54 [29] | |  | 71 [36] | 261 [28] | | 222 [21] | | | 54 [16] |
| BP meds | 156 [83] | 151 [88] | 192 [80] | | | | | | 132 [71] | |  | 127 [64] | 561 [59] | | 582 [54] | | | 158 [48] |
| Cholesterol meds, | 93 [49] | 87 [51] | 125 [53] | | | | | | 86 [47] | |  | 112 [57] | 507 [54] | | 496 [46] | | | 160 [48] |
| BMI, (kg/m^2^) | 30 (4.8) | 29 (4.9) | 29 (4.7) | | | | | | 29 (4.2) | |  | 29 (5.7) | 28 (5.2) | | | 28 (5.1) | | 28 (5) |
| SBP, mmHg | 135 (19) | 136 (19) | 134 (18.2) | | | | | | 131 (16) | |  | 131 (19.6) | 130 (17.4) | | | 129 (18) | | 127 (16.6) |
| DBP, mmHg | 70 (9.7) | 70 (10.6) | 70 (10.1) | | | | | | 70 (9.6) | |  | 64 (11) | 65 (10.4) | | 66 (10.4) | | | 66 (10.6) |
| HDL-C, mg/dl | 53 (13.5) | 55 (13.5) | 54 (13.6) | | | | | | 57 (14.3) | |  | 48 (12.2) | 52 (13.6) | | 53 (14.7) | | | 52 (13.8) |
| Total cholesterol, mg/dl | 186 (38) | 186 (40.2) | 187 (44.2) | | | | | | 189 (36.1) | |  | 176 (42.9) | 182 (43) | | 180 (42.4) | | | 180 (40.5) |
| cf-PWV, m/s | 13 (3.7) | 12.4 (3.5) | 12.3 (3.5) | | | | | | 11.4 (3.2) | |  | 12.1 (3.2) | 12 (4.6) | | 11.4 (3.4) | | | 11.1 (3) |

Mean(SD) or N[%]; **Abbreviations:** ARIC-Atherosclerosis Risk In Communities; HS-High School; NC-North Carolina, MS-Mississippi, MN-Minnesota, MD-Maryland; BMI-body mass index; SBP-Systolic blood pressure; DBP-diastolic blood pressure; HDL-C-high density lipoprotein cholesterol; cf-PWV-central femoral pulse wave velocity;

**Table 2. Characteristics of study participants by Income and Race, ARIC, 2011-2013**

|  | **African American (N = 789)** | | | | | |  | | **White (N = 2573)** | | | | | | | | | | | | | | | | | |
| --- | --- | --- | --- | --- | --- | --- | --- | --- | --- | --- | --- | --- | --- | --- | --- | --- | --- | --- | --- | --- | --- | --- | --- | --- | --- | --- |
| M(SD) or N [%] | **<$25K**  396 [50] | | | | **$25K-<$50K**  196 [25] | **≥$50K**  197 [25] | |  | | **<$25K**  469 [18] | | | | | | | **$25K-<$50K**  910 [35] | | | | | | | | | **≥$50K**  1194 [46] |
| Age, yrs | | | 75 (4.9) | 73 (4.4) | | 73 (4.6) | |  | | | 77 (5.2) | | | | | 75 (4.9) | | | | | | 74 (4.6) | | | | |
| **Sex** | | |  | | |  |  |  |  |  |  | | | | |  | | | | |  |  | | | | |
| Females | | | 303 [77] | 141 [72] | | 108 [55] | |  | | | 353 [72] | | | | | 593 [65] | | | | | | 638 [53] | | | | |
| **Site** | | |  | | |  | |  | | |  | |  | | | | | | |  | | | | | | |
| Forsyth Co., NC | | | 17 [4] | 21 [11] | | 7 [4] | |  | | | 91 [19] | | | | | 206 [23] | | | | | | 332 [28] | | | | |
| Jackson, MS | | | 379 [96] | 175 [89] | | 190 [96] | |  | | | - | | | | - | | | | | | | - | | | | |
| Minneapolis suburbs, MN | | | - | - | | - | |  | | | | 134 [29] | | | | 349 [38] | | | | | | 569 [48] | | | | |
| Washington Co., MD | | | - | - | | - | |  | | | | 244 [52] | | | | 355 [39] | | | | | | 293 [25] | | | | |
| **Education** | |  | | | |  | |  |  | | | | | | | | | | |  | | | | |  | |
| <HS | | 156 [40] | | 23 [12] | | 12 [6] | |  | | | | 105 [22] | | | | | | 65 [7] | | | | | | | 28 [2] | |
| HS | | 117 [30] | | 44 [22] | | 12 [6] | |  | | | | 220 [47] | | | | | | 417 [46] | | | | | | | 313 [26] | |
| Some college or college | | 104 [26] | | 70 [36] | | 65 [33] | |  | | | | 134 [29] | | | | | | 368 [40] | | | | | | | 589 [49] | |
| Post-graduate | | 19 [5] | | 59 [30] | | 108 [55] | |  | | | | 10 [2] | | | | | | 60 [7] | | | | | | | 264 [22] | |
| **Lifestyle** | |  | |  | |  | |  | | | |  | |  | | | | | |  | | | | |  | |
| Current alcohol | | 64 [16] | | 45 [23] | | 68 [35] | |  | | | | 199 [42] | | | | | | 500 [55] | | | | | | | 872 [73] | |
| Current smoker | | 31 [8] | | 10 [5] | | 7 [4] | |  | | | | 36 [8] | | | | | | 38 [4] | | | | | | | 75 [6] | |
| **CV risk factors** | |  | |  | |  | |  | | | |  | |  | | | | | |  | | | | |  | |
| Diabetes | | 173 [44] | | 71 [36] | | 68 [35] | |  | | | | 150 [32] | | | | | | | 216 [24] | | | | | | 242 [20] | |
| BP meds | | 333 [85] | | 161 [82] | | 137 [70] | |  | | | | 272 [58] | | | | | | | 533 [59] | | | | | | 623 [53] | |
| Cholesterol meds | | 194 [49] | | 100 [52] | | 97 [50] | |  | | | | 240 [51] | | | | | | | 451 [50] | | | | | | 584 [49] | |
| BMI, (kg/m^2^) | | 29.5 (4.9) | | 29 (4.3) | | 29 (4.5) | |  | | | | 28 (4.7) | | | | | | | 27 (4.3) | | | | | 27 (4.2) | | |
| SBP, mmHg | | 135 (19.4) | | 133 (18.1) | | 132 (15.5) | |  | | | | 131 (18.7) | | | | | | | 130 (17.5) | | | | | 127 (16.2) | | |
| DBP, mmHg | | 69 (10.2) | | 70 (9.8) | | 71 (9.6) | |  | | | | 64 (10.2) | | | | | | | 66 (9.8) | | | | | 66 (10.2) | | |
| Total cholesterol, mg/dl | | 186 (39.2) | | 191 (43.1) | | 185 (38.2) | |  | | | | 52 (13.5) | | | | | | | 54 (14.2) | | | | | 54 (13.8) | | |
| HDL-C, mg/dl | | 54 (13.3) | | 56 (14.7) | | 54 (13.5) | |  | | | | 190 (41.4) | | | | | | | 190 (41.5) | | | | | 187 (39) | | |
| cf-PWV, m/s | | 13.0 (3.7) | | 12.0 (3.2) | | 12.0 (3.2) | |  | | | | 12.2 (4.3) | | | | | | | 12.0 (3.7) | | | | 11.1 (3.8) | | | |

Mean(SD) or N[%]; **Income:** K denotes $1000 USD; **Abbreviations:** ARIC-Atherosclerosis Risk In Communities; NC-North Carolina, MS-Mississippi, MN-Minnesota, MD-Maryland; BMI-body mass index; SBP-Systolic blood pressure; DBP-diastolic blood pressure; HDL-high density lipoprotein; cf-PWV-central femoral pulse wave velocity; hs-CRP-high-sensitivity c-reactive protein.

Table 3. Association of Socioeconomic Status with Carotid Femoral Pulse Wave Velocity, ARIC, 2011-2013 Overall N=**3362**.

|  | **Model 1**  **N=3362** | | | | | | | **Model 2**  **N=3307** | | | | | | | | | **Model 3**  **N=3290** | | | | |
| --- | --- | --- | --- | --- | --- | --- | --- | --- | --- | --- | --- | --- | --- | --- | --- | --- | --- | --- | --- | --- | --- |
|  | ***β*** | | | | ***95% CI*** | | ***p*-value** | | ***β*** | | | | **95% CI** | | ***p-*value** | | | ***β*** | | **95% CI** | ***p-*value** |
| Education |  | | | | |  | | | | | | | |  |  |  |  |  |  |  |  |
| <HS | Ref |  | | | | |  | | | |  | | |  | |  |  | |  | |  |
| HS | -0.03 | | -0.47, 0.42 | | | | 0.90 | | | 0.03 | | | | -0.42, 0.48 | | 0.90 | 0.03 | | -0.40, 0.47 | | 0.88 |
| ≥Some college | -0.29 | | -0.73, 0.16 | | | | 0.21 | | | -0.10 | | | | -0.55, 0.35 | | 0.65 | -0.05 | | -0.49, 0.39 | | 0.82 |
| Post-graduate | -0.94 | | -1.44, -0.44 | | | | <0.001 | | | -0.63 | | | | -1.14, -0.12 | | 0.02 | -0.45 | | -0.95, 0.04 | | 0.07 |
| Income  <$25K | Ref | | |  | | |  | |  | | |  | | | |  |  | |  | |  |
| $25K-<$50K | -0.56 | | | -0.91, -0.22 | | | 0.002 | | -0.45 | | | -0.80, -0.10 | | | | 0.01 | -0.45 | | -0.78, -0.11 | | 0.01 |
| $50K ≥ | -0.82 | | | -1.16, -0.47 | | | <0.001 | | -0.64 | | | -1.00, -0.29 | | | | <0.001 | -0.53 | | -0.88, -0.19 | | 0.002 |

Model 1-linear regression model adjusted for age, site, sex, race; Model 2-Model 1 + CV risk factors (HDL, total cholesterol, BMI, smoking, alcohol), lipid lowering medications, diabetes; Model 3-Model 2 + BP, BP medications
