## Supplementary Tables 1, 2, 3 for "Association of socioeconomic status with arterial stiffness in older African American and White Adults: The ARIC Study Cohort"

Supplementary Table 1. Association of Socioeconomic Status and Carotid Femoral Pulse Wave Velocity in African American Adults, ARIC Study, 2011-2013.

**African Americans**

|  | Model 1  *n*=789 | | | | Model 2  *n*=765 | | | | | | | | | Model 3  *n*=763 | | | | | |
| --- | --- | --- | --- | --- | --- | --- | --- | --- | --- | --- | --- | --- | --- | --- | --- | --- | --- | --- | --- |
|  | ***β*** | ***95% CI*** | *p*-value | | | | ***β*** | | | ***95% CI*** | *p*-value | | | | ***β*** | ***95% CI*** | | | *p*-value |
| Education |  | | | |  | | | | | | | | | |  | | | | |
| <HS | Ref | | | | - | | | | | | | | | | - | | | | |
| HS | -0.40 | -1.10, 0.30 | | 0.26 | | | -0.44 | | -1.15, 0.26 | | | 0.21 | -0.62 | | | | | -1.28, 0.05 | 0.07 |
| ≥Some college | -0.23 | -0.88, 0.43 | | 0.50 | | | -0.20 | | -0.87, 0.46 | | | 0.55 | -0.26 | | | | | -0.89, 0.38 | 0.43 |
| Post-graduate | -1.29 | -1.98, -0.60 | | <0.001 | | | -1.14 | | -1.86, -0.43 | | | 0.002 | -0.97 | | | | | -1.65, -0.29 | 0.001 |
| Income |  | | | | |  | | | | | | |  | | | | | | |
| <$25K (ref) | - | | | | | - | | | | | | | - | | | | | | |
| $25K-<$50K | -0.90 | -1.49, -0.31 | 0.002 | | | -0.79 | | -1.39, -0.19 | | | | 0.01 | -0.69 | | | | -1.26, -0.13 | | 0.02 |
| $50K ≥ | -0.86 | -1.46, -0.27 | 0.005 | | | -0.74 | | -1.35, -0.13 | | | | 0.02 | -0.57 | | | | -1.15, 0.01 | | 0.05 |

Model 1- linear regression model adjusted for age, site, sex; Model 2-Model 1 + CV risk factors (HDL, total cholesterol, BMI, smoking, alcohol), lipid lowering medications, diabetes; Model 3-Model 2 + BP, BP medications.

Supplementary Table 2. Associations of Socioeconomic status and Carotid Femoral Pulse Wave Velocity in Whites,

ARIC Study, 2011-2013.

|  | | | | Model 1  *n*=2573 | | | | | | | | Model 2  *n*=2540 | | | | | | | | | | | Model 3  *n*=2525 | | | | | | | | | |
| --- | --- | --- | --- | --- | --- | --- | --- | --- | --- | --- | --- | --- | --- | --- | --- | --- | --- | --- | --- | --- | --- | --- | --- | --- | --- | --- | --- | --- | --- | --- | --- | --- |
|  | | | | ***β*** | ***95% CI*** | | | *p*-value | | | ***β*** | | | | | ***95% CI*** | | | *p*-value | | | | | | ***β*** | | | ***95% CI*** | | | | *p*-value |
| Education |  | | | | | | | | | |  | | | | | | | | | | |  | | | | | | | | | | |
| <HS | | Ref | | | | | | | | | - | | | | | | | | | | | - | | | | | | | | | | |
| HS | | | 0.16 | | | | -0.43, 0.76 | | | 0.58 | 0.30 | | | | -0.30, 0.89 | | | | | 0.32 | | | | | 0.39 | -0.19, 0.97 | | | | 0.18 | | |
| ≥Some college | | | -0.17 | | | -0.78, 0.43 | | | | 0.57 | 0.11 | | | -0.50, 0.72 | | | | | | 0.72 | | | | | 0.25 | | -0.35, 0.84 | | | | 0.42 | |
| Post-graduate | | | -0.68 | | | -1.38, 0.01 | | | | 0.05 | -0.28 | | | -1.00, 0.43 | | | | | | 0.43 | | | | | -0.06 | | -0.76, 0.63 | | | | 0.86 | |
| Income^*^ |  | | | | | | | | | |  | | | | | | | | | | |  | | | | | | | | | | |
| <$25K (ref) | Ref | | | | - | | | | | | - | | | | | | - | | | | | - | | | | | | | - | | | |
| $25K-<$50K | -0.42 | | | | -0.85, 0.01 | | | | 0.05 | | -0.32 | | -0.75, 0.11 | | | | | | | | 0.15 | | | -0.35 | | | | -0.77, 0.07 | | 0.10 | | |
| $≥$50K | -0.74 | | | | -1.17, -0.31 | | | | 0.001 | | -0.55 | | -0.99, -0.11 | | | | | | | | 0.01 | | | -0.48 | | | | -0.90, -0.05 | | 0.03 | | |

Model 1- linear regression model adjusted for age, site, sex; Model 2-Model 1 + CV risk factors (HDL, total cholesterol, BMI, smoking, alcohol), lipid lowering medications, diabetes; Model 3-Model 2 + BP, BP medications.

Supplementary Table 3. Main and Interaction effects between education, income, and race on PWV, ARIC Study, 2011-2013

|  | **Model 1**  **N=3362** | | | | | | | | **Model 2**  **N=3307** | | | | | | | **Model 3**  **N=3290** | | | | |
| --- | --- | --- | --- | --- | --- | --- | --- | --- | --- | --- | --- | --- | --- | --- | --- | --- | --- | --- | --- | --- |
|  | ***β*** | | | | | ***95% CI*** | | ***p*-value** | | ***β*** | | **95% CI** | | ***p-*value** | | | ***β*** | | **95% CI** | ***p-*value** |
| Education |  | | | | | |  | | | | | |  |  |  |  |  |  |  |  |
| <HS | | | 1.30 | 0.55, 2.05 | | | | 0.001 | | 1.07 | | | 0.32, 1.82 | | 0.01 | 0.94 | | 0.21, 1.67 | | 0.01 |
| HS | | | 0.86 | 0.09, 1.63 | | | | 0.03 | | 0.60 | | | -0.17, 1.38 | | 0.13 | 0.30 | | -0.45, 1.05 | | 0.44 |
| ≥Some college | | | 1.01 | 0.30, 1.72 | | | | 0.01 | | 0.83 | | | 0.11, 1.55 | | 0.02 | 0.65 | | -0.04, 1.35 | | 0.07 |
| Post-graduate | | | Ref | | | | | | | - | | | | | | - | | | | |
| **Interaction** |  | | | |  | | |  | |  |  | | | |  |  | |  | |  |
| Education X race | |  | | |  | | | 0.29 | |  |  | | | | 0.27 |  | |  | | 0.11 |
| Income  <$25K | | 0.74 | | | 0.10, 1.38 | | | 0.02 | | 0.68 | 0.04, 1.32 | | | | 0.04 | 0.50 | | -0.12, 1.12 | | 0.12 |
| $25K-<$50K | | -0.20 | | | -0.94, 0.53 | | | 0.59 | | -0.14 | -0.88, 0.60 | | | | 0.71 | -0.25 | | -0.96, 0.47 | | 0.50 |
| $50K ≥ | | ref | | |  | | |  | |  |  | | | |  |  | |  | |  |
| **Interaction** |  | | | |  | | |  | |  |  | | | |  |  | |  | |  |
| Income X race | |  | | |  | | | 0.30 | |  |  | | | | 0.37 |  | |  | | 0.46 |

Model 1-linear regression model adjusted for age, site, sex; Model 2-Model 1 + CV risk factors (HDL, total cholesterol, BMI, smoking, alcohol), lipid lowering medications, diabetes; Model 3-Model 2 + BP, BP medications.
